# Basic Baseline model design choices can substantially influence performance in collaborative forecast hubs

**DOI:** 10.64898/2026.03.18.26348748

**Authors:** Ehsan Suez, Spencer J. Fox

## Abstract

Over the past decade, outbreak forecasting has become an increasingly used tool to assist public health decision-making during epidemics. Collaborative forecast hubs, where multiple teams submit predictions in real-time, are the gold standard for such efforts. For each hub, a Baseline model is used as a performance benchmark for other models. Although the Baseline is understood as a naïve forecast, its design is subjective, and the impact of model design decisions remains understudied. We evaluated how three Baseline specification decisions influence forecast performance on trend models that forecast based on historically observed dynamics: (1) the amount of historical data used for training, (2) whether the data are transformed, and (3) whether forecasts follow a *flatline* variant (constant predictions) or a *drift* variant (allowing a slope). Retrospective forecasts were generated for multiple years across four surveillance targets: COVID-19, influenza and RSV hospital admissions, and weighted influenza-like illness percentage. For wILI, we additionally compared trend baselines with a seasonal baseline model leveraging long-term historical patterns. Model specification significantly altered performance. The optimal performing model across targets was a flatline model that used the most recent 6-12 transformed observations. The optimal model outperforms the current standard Baseline used in many forecast hubs by an average of 9.6% (range: 3.7-12.9%) across forecast targets, and it outperformed the seasonal baseline model by 32.3% across nine influenza seasons. Our results demonstrate that subjective Baseline design decisions can materially influence forecast accuracy and, consequently, the perceived rankings of models within collaborative forecast hubs. Based on the varying approaches and their performance differences, these findings highlight the need for increased transparency in Baseline model specifications and support the routine inclusion of multiple benchmark models within collaborative forecast hubs.

**Author Summary:** Forecasting the spread of infectious diseases is increasingly used to support public health decision-making during outbreaks. Many large forecasting efforts operate through collaborative forecast hubs, where multiple research teams submit predictions that are evaluated together. In these systems, a simple Baseline model serves as a benchmark against which more complex models are compared. However, the design of these Baseline models often involves subjective choices that have rarely been systematically studied. We examined how several common design decisions affect the performance of widely used Baseline approaches using multiple years of data for several respiratory diseases. We found that small changes in parameterization can substantially alter forecast accuracy and, consequently, the relative rankings of other models. Across all datasets, a single simple Baseline configuration consistently achieved the strongest performance, outperforming the most commonly used model. Our findings highlight the central and understudied role that Baseline models play in collaborative forecast hubs, and provide guidance to hub organizers regarding the appropriate design and communication of Baseline models.

## Introduction

Over the past few decades, the world has experienced outbreaks ranging from severe regional epidemics to global pandemics, alongside recurring seasonal diseases, all of which have produced substantial health, social, and economic consequences [1–5]. Infectious disease modeling and forecasting now play vital roles in responding to these outbreaks, helping to transform complex epidemiological data into actionable insights for decision-makers [6,7]. By anticipating the patterns of epidemiological burden indicators such as case counts or hospital admissions, forecasts support effective real-time decisions such as the optimal allocation of antiviral or vaccine resources or type and timing of transmission interventions [6,7].

In recent years, collaborative forecasting hubs have become the dominant paradigm for outbreak forecasting [8–10]. Collaborative hubs have been established in various forms over the years [11–13], playing a pivotal role in managing outbreaks of COVID-19, dengue, influenza, and Zika [9,10,14,15]. These hubs bring together multiple modeling teams to generate probabilistic forecasts for shared targets using diverse methodologies. Hub organizers then evaluate and combine these forecasts, often via ensemble models that serve as a single forecast for communication and tend to outperform any individual forecast model. A central component of a forecast hub is the Baseline model, which serves as a reference benchmark against which all other models are compared [9,10]. Since forecasts are commonly judged by their ability to outperform the Baseline model, the specification and performance of the Baseline are of central importance.

Baseline models vary widely in form. At the simplest end of the spectrum are so-called naïve Baseline models, which include *trend* and *seasonal* approaches. Trend models extrapolate observed dynamics forward based on changes between successive observations, whereas seasonal models generate forecasts using patterns observed in prior seasons. Within trend models, *flatline* predictors produce constant forecasts centered on the most recent observation, while *drift* predictors allow for non-zero slopes. In both flatline and drift variants, forecast uncertainty is derived from historical inter-observation variability. More complex approaches include models such as ARIMA (with or without covariates), exponential smoothing, neural networks, and many others [16–20]. Flatline predictors are the most commonly used benchmark, being used by the U.S. and European forecast hubs for influenza, COVID-19, and RSV as of 2026 [9,10,21–26]. Seasonal baseline models are commonly used for diseases with clear seasonal trends like influenza and dengue and were previously the Baseline model for U.S. FluSight challenges [8,11,27,28]. More complex Baseline models have been deployed for a wide range of pathogens including emerging and seasonal ones with established predictors (e.g., digital surveillance or climate indicators), such as for Ebola, dengue, and Zika [18,29–31].

Although more complex Baseline models are often presumed to yield better forecasts, evidence from collaborative forecast hubs and recent evaluations indicates that greater model complexity does not guarantee improved performance [8–10,32]. Despite their central role in hubs and the uncertainty in how their design may impact forecast performance, there has been limited systematic evaluation of the diversity of Baseline modeling approaches across diseases and contexts. Recently, Stapper and Funk proposed a structured framework for Baseline model selection and conducted a broad comparative analysis of Baseline models spanning a range of complexity [32]. Their work quantified important differences in Baseline performance and demonstrated how Baseline choice can influence the downstream relative rankings of forecast models for influenza and COVID-19. Notably, their findings highlighted that no single Baseline model was globally optimal, suggesting that collaborative forecast hubs may benefit from adopting context-specific Baseline models.

In this study, we carry out complementary work. Rather than comparing the full range of available Baseline model classes, we focus on variations within the simplest and most widely used approaches: trend and seasonal ones. This emphasis reflects the practical challenges faced by many forecast hub organizers, who may lack the data, time, or specialized expertise required to evaluate a broad set of candidate models before settling on one. It is also grounded in the original conceptualization of Baseline models as neutral and broadly applicable benchmarks, intended to provide robust, interpretable, and consistent performance for comparison rather than to serve as steep thresholds for competing models to surpass [8,28]. Even simple trend- and seasonal-based approaches require subjective choices, and the performance implications of these seemingly minor decisions remains poorly understood. Identifying simple, portable Baseline specifications that perform reliably across diseases and surveillance contexts could therefore offer substantial practical value to hub organizers.

Motivated by these considerations, we systematically evaluated variations of a widely used trend-based Baseline model to quantify how common specification choices influence forecast performance. Specifically, we assessed how data transformations and the amount of historical data used for model fitting affect the forecast accuracy of both drift and flatline trend models across COVID-19, influenza, RSV, and weighted influenza-like illness percentage (wILI). For wILI, which was the only target with sufficient long-term historical data, we additionally compared trend-based models with the standard seasonal Baseline model. Our results identify a Baseline configuration with optimal or near-optimal performance across diseases and datasets and provide practical guidance for collaborative forecast hub organizers selecting an appropriate Baseline model.

## Methodology

To guide Baseline model selection in collaborative forecasting hubs, we retrospectively compared the forecast performance of two classes of Baseline models commonly used in recent outbreak forecasting efforts: (1) trend baselines, which generate forecasts using recent time series dynamics, and (2) seasonal baselines, which generate forecasts using historically observed seasonal patterns. Within the trend-based class, we evaluated multiple model variants that differed only in key configuration choices, including the amount of historical data used for fitting, the application of variance-stabilizing transformations, and whether forecasts assumed persistence (*i*.*e*., flatline variants) or extrapolated recent trends (*i*.*e*., drift variants). These trend baseline variants were evaluated across multi-year time series for COVID-19 hospital admissions, influenza hospital admissions, RSV hospital admissions, and weighted influenza-like illness percentage (wILI). Seasonal baseline models were evaluated using wILI data only, as this outcome was the only one that provides a sufficiently long and stable historical record to support robust estimation of seasonal structure.

### Baseline model specification

#### Trend baseline models

We implemented the trend models using the quantile_baseline function made available in the simplets package in R that is currently used for FluSight and COVID-19 forecast hubs [33–35]. Briefly, we let *z*_*t*_ denote an observed value at time index *t* ∈ {1, …, *T*} within a time-series of length *T*. We explore whether a square root transformation that stabilizes the variance of the time-series impacts forecast performance. Specifically, we define 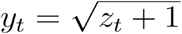 for transformed variants and *y*_*t*_ = *z*_*t*_ for non-transformed ones. We specify the forecast origin as *y*^∗^ = *y*_*T*_. Predictive distributions are generated using the empirical difference distribution defined as:

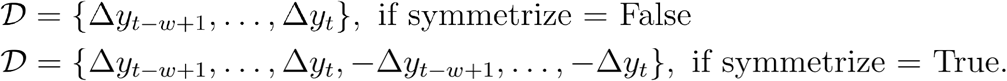

Where Δ*y*_*t*_ *= y*_*t*_ − *y*_*t* − 1_ for *t* ∈ {2, …, *T*}, *w* specifies the number of recent differences to use, and symmetrize = True is used for the *flatline* variant while symmetrize = False is used for the *drift* variant. Individual trend forecast trajectories are made by drawing monte carlo samples from the empirical difference distribution, 𝒟. For forecast horizon *h* ∈ {1, …, *H*} a simulated trajectory is defined as:

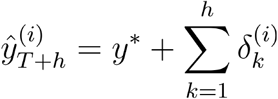

where *H* specifies the maximum desired forecast horizon, 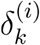 is the sampled difference for forecast horizon, *k*, randomly drawn from 𝒟, and *i* ∈ {1, …, *N*} indexes the *N* forecast trajectories. For the flatline variant (symmetrize=True), the trajectories are recentered so that the predictive medians at each horizon equal the forecast origin:

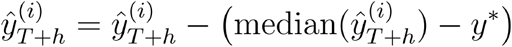

We back-transform the forecasts to the original scale as 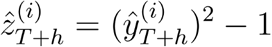 for transformed variants, and 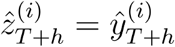 for untransformed variants. For count and percentage targets, forecasts are constrained to be non-negative: 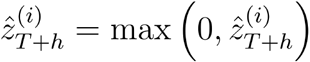 . For percentage targets, predictions are additionally allowed to have a maximum value of 1 as: 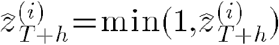 . We summarize the forecast trajectories for each horizon into 23 quantiles according to the recent COVID-19 hub, and FluSight forecast challenge specifications [36,37] as:

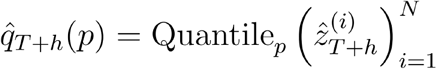

where 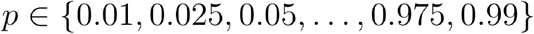.

#### Seasonal baseline model

As a model comparison, we implemented the same seasonal baseline used during the FluSight challenge from 2014-2018 and made available in code from [38]. Briefly, for the model we define *z*_*t*_ as the value of the time-series at time index *t* ∈ {1, …, *T*}. We model the time-series as a Generalized Additive Model (GAM) with a cyclic spline to capture annual patterns as:

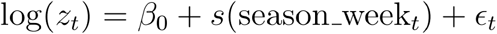

where *β*_0_ is the intercept term, *s*(season_week_*t*_) denotes a smooth cyclic spline evaluated at the season week corresponding to time, *t*, and Gaussian noise *ϵ*_*t*_. For each region forecasted, the seasonal baseline model was evaluated using a rolling-origin design, with the GAM fitted to the season’s available wILI data and used to generate 1-4-week-ahead forecasts. The evaluation period was aligned with that of the trend-based baseline models.

Probabilistic forecasts were generated by repeatedly simulating future outcomes from the fitted GAM. For each forecast date, we sampled forecast trajectories based on uncertainty in the estimated model coefficients. The simulated predictions were then transformed back to the original incidence scale, and forecast quantiles were obtained empirically from the resulting predictive distribution. To ensure valid probabilistic forecasts, we enforced that forecast quantiles were non-decreasing with quantile level, correcting rare numerical inconsistencies arising from finite simulation.

### Retrospective forecasting methodology

Following the methodologies from their respective hubs [21,23], we generated weekly retrospective forecasts in a rolling-origin, real-time–like manner for each model variant across four U.S. disease surveillance datasets. For each forecast date, models were trained using only data available up to that date.

For COVID-19 hospital admissions (November 2020–October 2023) and influenza hospital admissions (October 2023–May 2025), we used the surveillance data available at each forecast issue date, approximating real-time forecasting conditions. For COVID-19, we obtained the archived daily forecast hub snapshots from the versioned Johns Hopkins COVID-19 repository made available through the covidData R package [39] and subset to 145 weekly issue dates, for which we produced forecasts. For influenza we obtained weekly archived forecast hub snapshots from the FluSight GitHub repository [40]. In contrast to the revisions for influenza and COVID-19 hospital admissions, RSV hospital admissions (January 2022–April 2025) are subject to substantial reporting delays and large upward revisions (Fig S1). For RSV, we generated forecasts using both the weekly archived data and the finalized hospitalization count data as of January 2026 [40,41]. Most forecast hubs experience fewer revisions compared to the RSV data, so we present the results using the retrospective data in the main manuscript, but we present the results using the archived, real-time data in the supplement for hub organizers who anticipate large data revisions. For weighted influenza-like illness percentage (wILI; January 2014–April 2023), we additionally used the final values rather than the archived data as revisions were generally modest.

For influenza, RSV and COVID-19 hospital admissions, we generated forecasts for each U.S. state, Washington, D.C., Puerto Rico, and the national level, while we produced wILI forecasts for each of the ten Health and Human Services (HHS) regions and at the national level. For each target, we generated 1- to 4-week-ahead predictions weekly at the specific reference date. The influenza, RSV and wILI data were available at a weekly timescale, so the *t* in the time-series was weekly and *H* ∈ {1, …, 4} corresponds to the 1- to 4-week ahead predictions. For COVID-19, data are available at a daily scale, so the *t* in the time-series was daily and *H* ∈ {1, …, 28} corresponds to the 1- to 28-day ahead predictions. To standardize the forecast horizons, we consider the predictions for *H* ∈ {7, 14, 21, 28} as the one, two, three, and four week ahead forecasts respectively for COVID-19.

We explored how three factors impacted the forecast performance of the trend model: (1) the square root transformation, (2) the length of the window, *w*, used for the difference distribution, and (3) whether the flatline or drift variant was specified with the symmetrize parameter. For COVID-19, we considered training windows ranging from 5 to 360 daily observations, while for influenza hospital admissions, RSV, and wILI we considered windows ranging from 5 to 53 weekly observations; in all cases, we additionally evaluated a specification using the full historical record available up to each forecast reference date (*w* = ∞). We produced retrospective trend baseline forecasts for all combinations of model variants mentioned. For wILI, we additionally compared the trend forecasts with a seasonal baseline model that was fit in a rolling fashion, using all available historical data up to each forecast date, and evaluated over the same retrospective period.

### Forecast Evaluation Metrics

For all forecasts, performance was evaluated using the current standard metrics used in the forecast challenges, including the Weighted Interval Score (WIS) and Prediction interval coverage (PIC) [9,35,42]. The Weighted Interval Score (WIS) aggregates the accuracy of predictive intervals across multiple quantiles, taking into account the interval width and penalties for under- and over-prediction. For a set of nominal confidence levels *α*_1_, …, *α*_*K*_ and a predictive distribution *F*, let *l*_*k*_ and *u*_*k*_ denote the lower and upper bounds of the corresponding prediction intervals, and let *m* represent the predicted median. The WIS for an observed value *y* is then defined as

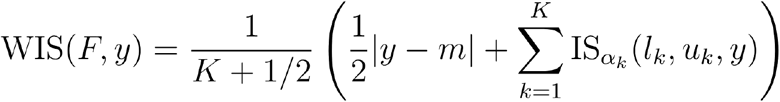

Where

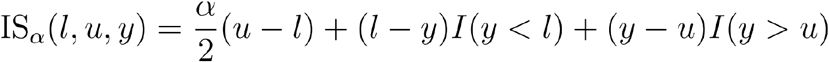

and I(·) is an indicator function that equals 1 when the condition is true and 0 otherwise [35]. Smaller WIS values indicate better forecast performance.

The Prediction Interval Coverage (PIC) for a given confidence level 1 − *α* equals 1 if the observed value falls within the corresponding prediction interval and 0 otherwise. Averaged over all forecasts, the PIC represents the proportion of observations captured by the (1 − *α*) × 100% interval [43]. Well-calibrated models yield PIC values close to 1 − *α*; here we assess 50% and 95% coverages. All forecast evaluation was performed using the scoringutils R package (version 1.2.2) [44].

## Results

Trend baseline models use recent observations and the inter-observation variation to generate forecasts, making their predictions sensitive to both their training data and model specification. As an illustrative example, we compare forecasts for COVID-19 hospital admissions issued on January 10, 2022, contrasting a flatline model, whose median forecast equals the most recent observed value, with a drift model, whose median reflects the estimated slope over the training window (Figure 1). When the most recent 10 days of data are used for model fitting, the median forecasts from the flatline and drift models diverge substantially. The drift model extrapolates the recent upward trajectory and predicts continued increases, whereas the flatline model projects a constant value equal to the most recent observation (Figure 1A). Correspondingly, the drift model produces prediction intervals that are shifted upward, while the flatline model yields symmetric intervals centered on the last observed value. Applying a data transformation yields similar median forecasts but results in asymmetric prediction intervals, with wider upper bounds and narrower lower bounds compared to the non-transformed model (Figure 1B). However, when all available data are used for model fitting, the median forecasts from the drift and flatline models become more similar, as periods of increase and decrease in the training data offset one another in the drift model (Figure 1C). A key difference also emerges in the prediction intervals, which are generally narrower when all available data are used for model fitting compared to when only recent data are used. This occurs because the full-data model averages inter-observation variability across the entire time series, whereas the recent-data model estimates variance based primarily on the larger, more volatile recent increases (Figure 1A vs. 1C).

**Figure 1:**
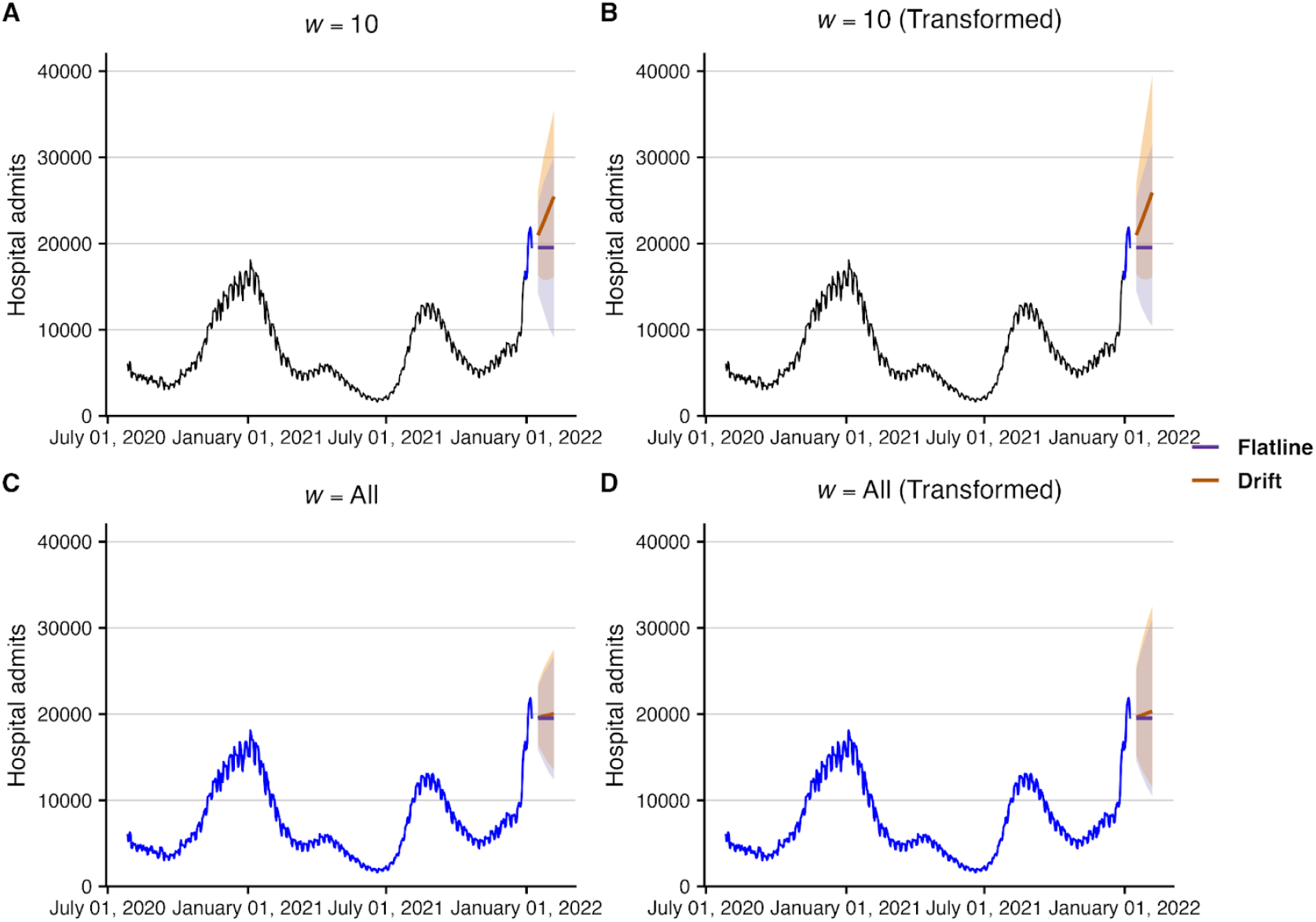
Impact of the model specification on predictions of trend baseline models using COVID-19 hospital admission data from July 1, 2020 to January 9, 2022 at the national level in the United States. Median (colored lines) and 95% prediction intervals (shaded ribbons) for 1-4 week ahead forecasts for the flatline (purple) and drift (orange) model variants under four training scenarios: ten days of raw data **(A)**, ten days of transformed data **(B)**, all historical data **(C)**, and all historical transformed data **(D)**. Observed daily hospital admissions are shown in black for data excluded from model fitting and in blue for data used in model training.

We first compared the performance of flatline and drift models using the most recent ten non-transformed observations across multiple years of COVID-19, influenza, and RSV hospital admissions, as well as weighted influenza-like illness percentage (wILI). In general, when models were fit using only recent data, drift variants performed best during periods of epidemic growth and decline, whereas flatline variants performed best near epidemic peaks and troughs (Figure 2; Figure S2). Across the full time series, the flatline model outperformed the drift model with mean WIS improvements of 10.5% for COVID-19, 8.3% for influenza, 10.4% for RSV, and 9.1% for wILI. The flatline model similarly outperformed the drift model across forecast horizons and geographies for each of the diseases (Figures S3-S4). A key reason that the flatline model outperforms the drift model may lie in its better coverage, with the 95% prediction interval being an average of 6.0% (range: 2.5-7.4%) higher across all four disease targets.

**Figure 2.**
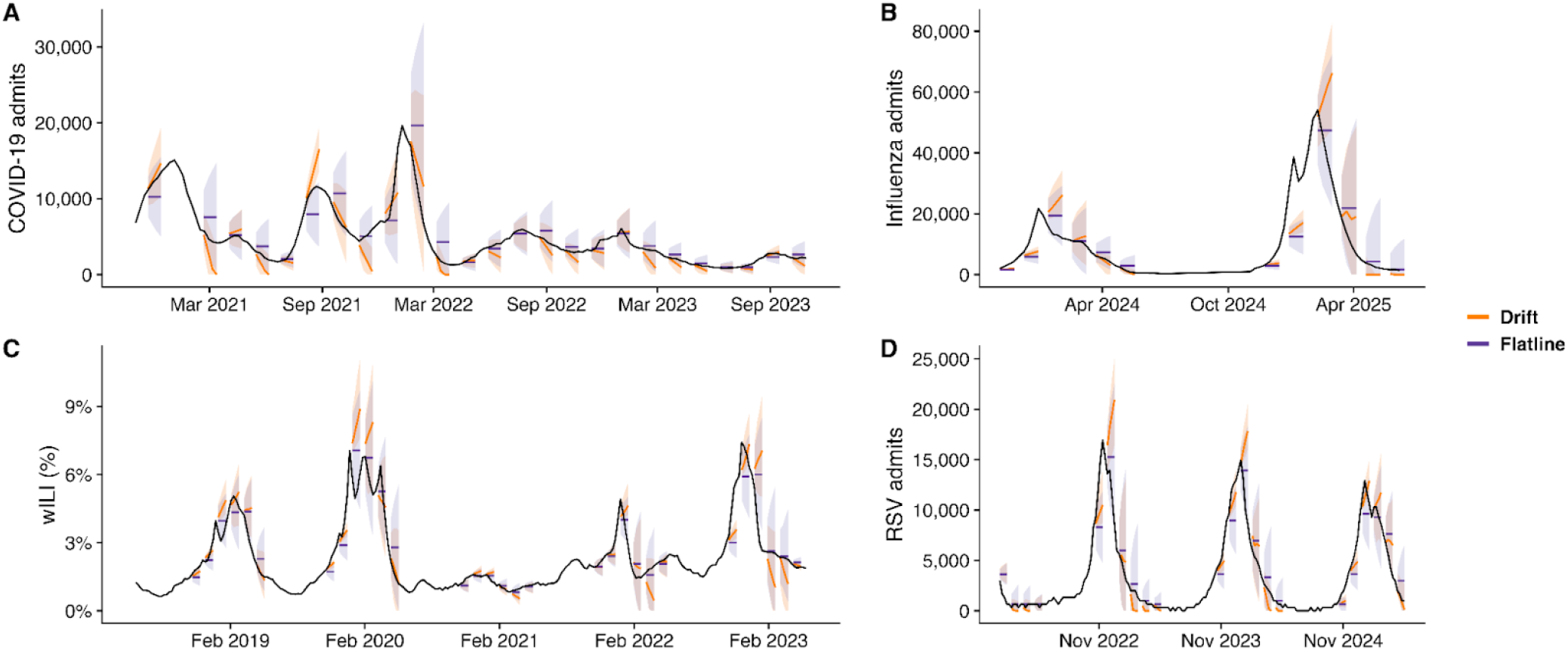
Retrospective forecasts for COVID-19, influenza, wILI, and RSV. Median (colored line) and 95% prediction intervals (shaded ribbons) for four week ahead forecasts from the flatline (purple) and drift (orange) baseline models that used 10 non-transformed observations for training. For COVID-19 and influenza, forecasts were generated each week using the data that were available in real-time, while forecasts were generated using the finalized data for wILI and RSV.

We investigated how forecast performance of each of the models varied as a function of the training data length (Figure 3). For drift baseline models, performance was optimal using fewer than the most recent 15 data points (COVID-19 and RSV) or for using all of the historical data (influenza and wILI). For flatline models, using fewer than 15 data points was the optimal training window for all diseases except for RSV, which had performance improvements of <1% when additional data were used (Figure 3D). Switching a model from using all of the available raw observations for training, which is the current standard Baseline used by collaborative forecast hubs, to using the most recent ten transformed observations improved forecast performance by an average of 9.6% (range: 3.7-12.9%) across all diseases (Table 1). When using the most recent ten observations, transforming the data improved the forecast performance by an average of 3.4% (range: -1.1%-6.2%) across all diseases (Table 1). Similar results were observed when comparing model performance by prediction interval coverage (PIC), with flatline models achieving PICs above those of the corresponding drift model (Figure S5-S6). Interestingly, non-transformed models had PICs closest to their nominal values for the 50% prediction interval, but the opposite was true for the 95% prediction intervals. Across targets, model PIC was generally optimized when fewer than 15 historical observations were used, with the exception of COVID-19.

**Table 1:**
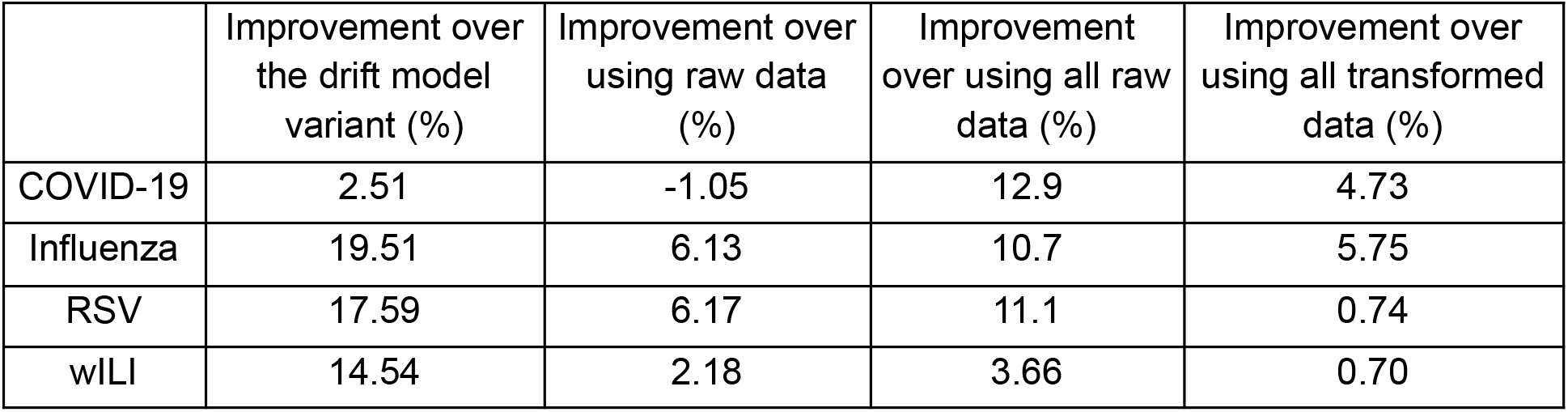
Performance improvement of the near-optimal forecast model, the flatline baseline model that uses ten transformed observations against variations. Forecast performance of the near-optimal model compared against the analogous drift model variant, the analogous flatline variant using raw observations, the flatline variant using all available raw observations, which is the current standard Baseline model used by most hubs, and the flatline variant using all available transformed observations.

**Figure 3:**
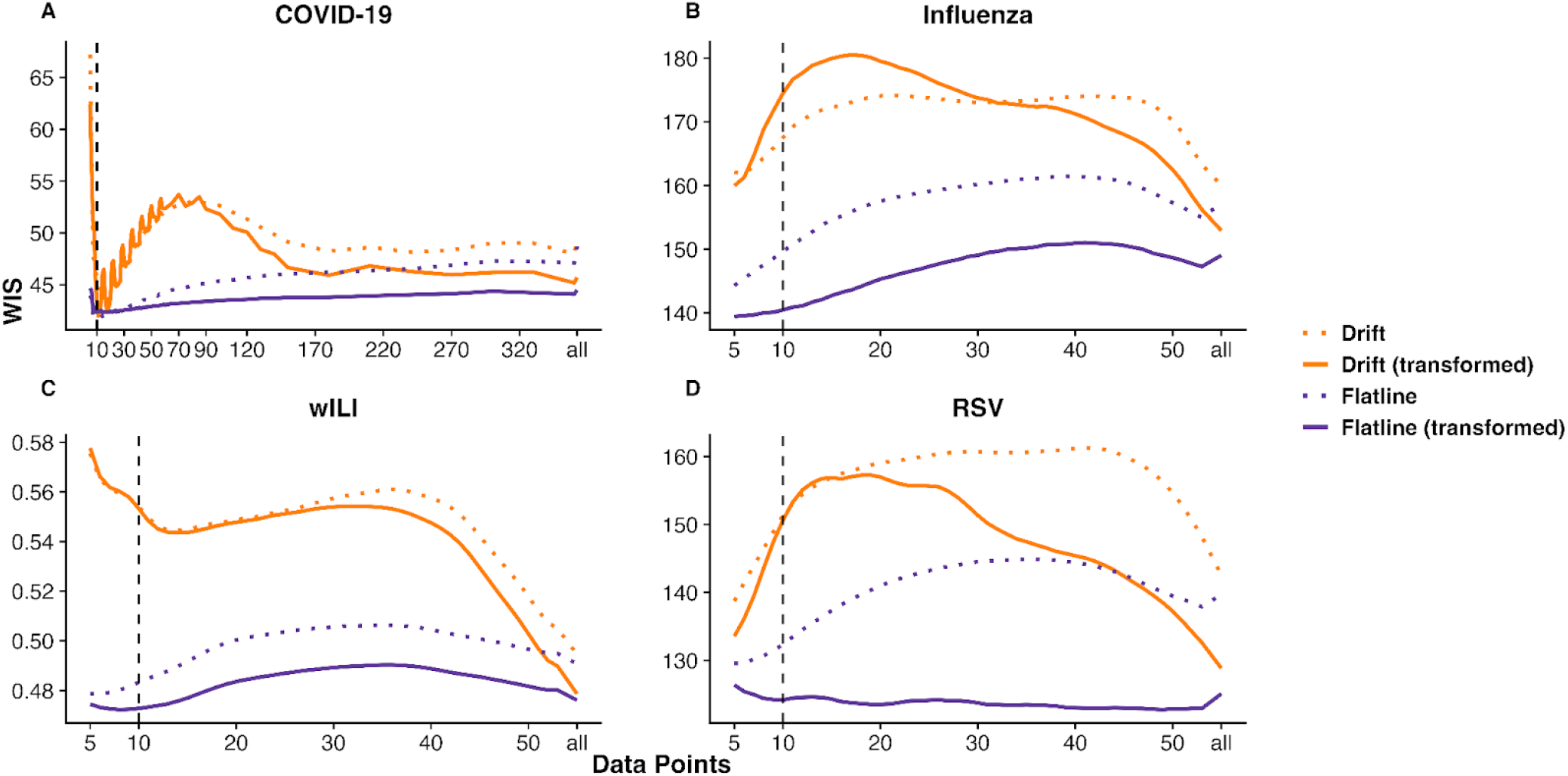
Performance of trend baseline model variants as a function of the training data window. One to four week ahead forecasts for each model variation were made weekly for all regions from November 2020 to October 2023 (COVID-19), October 2023 to May 2025 (Influenza), January 2014 to April 2023 (wILI), and January 2022 to April 2025 (RSV). Weighted Interval Score (WIS) estimates are summarized across forecast horizons, dates, and regions for COVID-19 **(A)**, Influenza **(B)**, wILI **(C)**, and RSV **(D)**. Colored lines indicate the WIS estimates for either the drift (Orange) or flatline (Purple) model variant using a specific training window and either transformed (solid lines) or raw (dotted lines) observations. The number of data points (x-axis) corresponds to days for COVID-19 and weeks for the other diseases, with a value of “all” indicating that all historical data prior to the forecast date are used for training. Lower WIS values indicate better forecast performance, and the vertical dashed line indicates the near-optimal model discussed in the main text and Table 1.

The optimal model configuration for RSV differed when forecasts were generated using archived, real-time data rather than retrospective data, underscoring the impact of reporting backfill on model performance (Figure S7A). When using real-time data, the drift model using 15 observations outperformed all other models, though it was only 0.3% better than the flatline model using all available data. Performance improvements of the drift model relative to the flatline variant increased as data revisions became larger (Figure S7B). Based on the observed forecast trajectories, the drift model outperformed the flatline model when using archived data in this case because of a unique dynamic where the largest revisions occurred near the epidemic peak. The drift model’s upward-sloping forecasts better matched the revised observations, whereas the flatline model was negatively biased during these time periods.

Finally, we compared four-week-ahead forecasts from the optimal trend model (*i*.*e*., the flatline model using ten transformed observations) with the historical FluSight hub seasonal baseline model for the wILI target across nine seasons from October 2014 through May 2023 (Figure 4A) [8,28]. Aggregated across all seasons, regions, and forecast horizons, the flatline model outperformed the seasonal model by 37.4% as measured by WIS. This advantage declined as a function of the forecast horizon, with the flatline model outperforming the seasonal model by 68.5%, 46.4%, 26.6%, and 8.5% for the one-, two-, three-, and four-week ahead forecast horizons, respectively (Figure S8). Across all nine seasons, the flatline model outperformed the seasonal baseline by an average of 37.1% (range: 7.5%-85.8%) per season and for 82.8% of forecast dates (Figures 4B-4C). Interestingly, the seasonal model tended to outperform the flatline model before and after the peak, though the flatline performed significantly better during the peak phase of the epidemic, a dynamic that was consistent across geographic regions (Figure 4B-4C). The flatline model outperformed the seasonal model despite exhibiting lower prediction interval coverage (PIC), with empirical coverage of 38.6% and 78.1% for the nominal 50% and 95% intervals, respectively, compared with 51.7% and 94.0% for the seasonal model.

**Figure 4:**
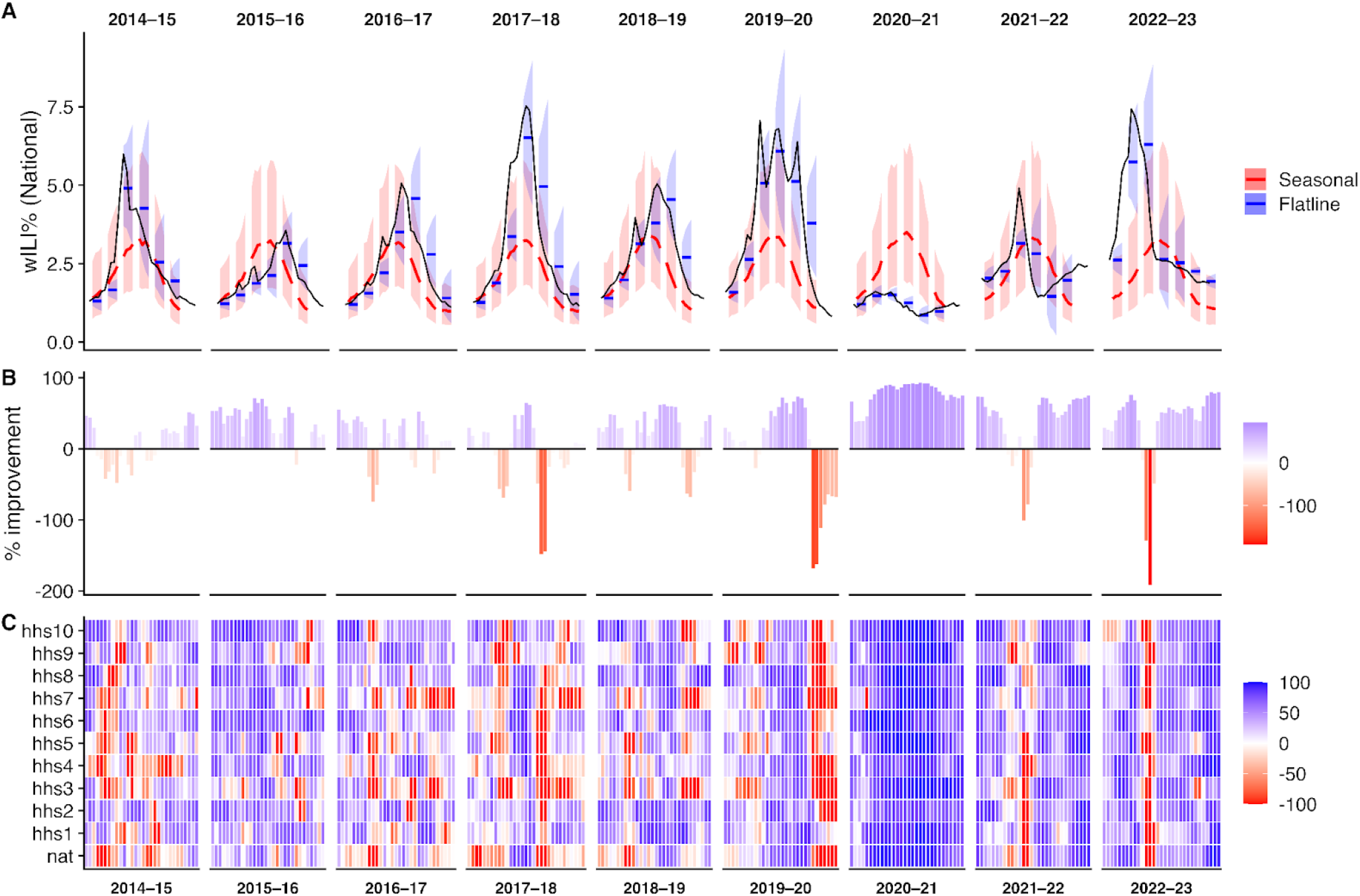
Comparative performance of the optimal trend and the seasonal baseline model for wILI from October 2014 to May 2023. **(A)** Retrospective four-week-ahead median (colored lines) and 95% prediction intervals (shaded ribbons) for forecasts for wILI at the National level from the seasonal (blue) and the ten-week transformed flatline model (red). **(B)** Percent improvement of the flatline model compared to the seasonal model averaged across all regions and forecast horizons for each of the 132 forecast dates. **(C)** Percent improvement of the flatline model compared to the seasonal model for each region averaged across all forecast horizons for each of the 132 forecast dates. For panels B and C, the color indicates the relative performance of the models, with blue indicating that the flatline model outperformed the seasonal model and red indicating the seasonal model outperformed the flatline model.

## Discussion

The field of outbreak forecasting has expanded significantly since the first influenza forecast challenge organized by the CDC in 2013 [45]. While Baseline models, which benchmark other models’ performance, were not part of the first competition, they soon took on a central role: providing a performance threshold for other forecast models to overcome [8,28]. Despite their importance, there has been no comprehensive analysis of how subjective design choices within the simplest and most commonly used Baseline models impact their performance and, consequently, the assessment of competing forecasting models. We systematically evaluated how the training data and model specifications of trend forecast models impact their performance across four multi-year datasets, demonstrating performance differences between model variants exceeding 40% (Figure 2). Our results identified a single Baseline configuration with near-optimal performance across all datasets: a flatline model trained on the ten most recent transformed observations, which achieved approximately a 10% improvement over the current standard Baseline model (Table 1). Taken together, our results provide clear guidance for forecast hub organizers in selecting a Baseline model and underscore the need for greater transparency in Baseline methodologies to ensure proper interpretation of existing and future hub results.

Our analysis highlights several technical insights about simple Baseline models. First, we find that trend models that emphasize recent observations outperform seasonal models, even for highly seasonal diseases like influenza (Figure 4). These findings support the recent move away from seasonal baselines towards flatline baselines in recent forecasting challenges for improving Baseline model performance [21,23,24]. However, our results suggest that as the forecast horizon increases, the performance between the two model types converges (Figure S8). Second, and similar to previous forecasting work, our results suggest that data transformations regularly improve forecast performance, presenting an opportunity to improve the performance of existing Baseline models for current forecasting hubs (Figure 3). These dynamics may stem from improved 95% prediction interval coverage of models built using transformed data (Figure S5-S6). Third, we observed that flatline and drift models converge in performance as the amount of historical data used for fitting increases. This pattern is intuitive, as incorporating longer time series captures both periods of increase and decrease, causing the estimated drift to average toward zero and thereby approximate the flatline model (Figure 1).

While Baseline models have been used in every recent forecast challenge, there is no consensus definition of what makes a good Baseline model in the literature: should the Baseline serve as a strong, challenging benchmark that only the best models can surpass, or should the Baseline be built upon salient features in the underlying data (e.g., seasonal dynamics) regardless of the ultimate performance? Consistent with the original conceptualization of Baseline models and our experience working with public health officials, Baseline forecasts are generally expected to function as neutral reference points—approximating what an informed observer might reasonably project [8,28,46]. In the simplest cases, such an observer may make predictions based on observed week-over-week variation, the slope of recent dynamics, or historical seasonality, which correspond to the flatline, drift, and seasonal models from our analysis, respectively. Our findings demonstrate that even simple Baseline models can exhibit substantial performance variability and the optimal model outperforms the current standard by 10%, with meaningful implications for the comparative evaluation of forecasting models. For example, a 10% improvement in performance of the Baseline model would reduce the proportion of models outperforming it by 14.9% (range: 0-30.4%) across recent COVIDHub and FluSight hub seasonal efforts in the U.S. (Figure S9).

Recent work by Stapper and Funk has enumerated five criteria for designing Baseline models, including: (1) capturing fundamental epidemiological characteristics (*e*.*g*., exponential growth or seasonality); (2) producing forecasts in the required format; (3) relying only on training data available to all competing models; (4) maintaining simplicity; and (5) ensuring that uncertainty intervals are well calibrated both overall and across forecast strata (*e*.*g*., locations) [32]. These criteria encompass a broader range of Baseline models than those examined in our study, but their findings are consistent with ours in demonstrating that Baseline model choice can substantially influence performance. Rather than emphasizing the breadth of model complexity they investigated, we focused on understanding the behavior and sensitivity of the trend and seasonal Baseline models most commonly used to benchmark performance in recent forecasting challenges. By focusing on simple and robust Baseline models, we aim to provide practical, broadly generalizable guidance for collaborative forecast hubs operating across diverse regions, diseases, and populations.

In that vein, our results offer clear direction for future forecast hub organizers: a flatline model trained on the most recent ten transformed observations achieved the strongest or nearly the strongest overall performance across the datasets examined (Figure 3). This configuration likely surpasses the commonly used flatline model, which relies on all available historical data, as it better captures the variance associated with the current epidemic phase, resulting in better-calibrated 95% prediction interval coverage compared to other models (Figure S6). A key aspect of this may be increasing anticipated inter-observation variability during the rise and decline of epidemic waves, when variability is highest. Drift models that follow recent trends may serve as a good naive-prediction benchmark, as they provide natural expectations for dynamic time series; however, they perform poorly because they tend to overshoot epidemiological peaks (Figure 2; Figure S2). Seasonal baseline models also yield reasonable benchmarks, but they lack the ability to recalibrate forecasts based on recent data and thus perform worse overall than the trend models, particularly during anomalously small or large seasons (Figure 4). Because forecasting hubs differ in their goals and contexts, there may be no global optimal Baseline for all efforts. However, our findings alongside previous results suggest that collaborative forecast hubs would likely benefit from adopting multiple Baseline models to benchmark performance [32]. For example, they may include an optimal flatline model as a stringent benchmark, a flatline model trained on all historical data for robust performance, and a seasonal baseline to test whether information from the current season improves predictions. One of these models can be chosen as the overall benchmark, but others can be used to further evaluate the performance of competing models.

Despite the central role that Baseline models play in collaborative forecast hubs, their configurations are not always described with the same level of detail as submitted forecasting models, and updates over time may not be systematically documented. While several hubs make Baseline model code publicly available (e.g. the FluSight hub [47]), formal reporting of configuration choices and their evolution is often limited in manuscripts and hub documentation. Hubs could further strengthen transparency by maintaining version-controlled Baseline code and explicitly reporting configuration decisions and updates over time. Such practices would enhance reproducibility, support fair model comparison, and reinforce confidence in collaborative forecasting efforts.

Our analysis has several limitations that may affect the interpretation and generalizability of our findings. First, we focused on respiratory disease dynamics in the United States, all of which exhibit at least partial seasonality [43]. Nonetheless, there are reasons to expect that our results may extend to a broader range of infectious disease contexts. COVID-19 displayed comparatively inconsistent seasonal structure during the study period, suggesting that our findings may be informative for emerging infectious diseases with less predictable seasonality. For transmission systems with stronger direct climate drivers, Baseline models that incorporate weather or environmental covariates may be appropriate and warrant further evaluation [11,20,32,48]. Second, we conducted simulated real-time forecasts only for influenza and COVID-19, with our results for wILI and RSV being based on retrospective data. Our findings are likely to generalize to diseases with relatively stable reporting dynamics. However, sensitivity analyses using the real-time RSV data that is characterized by substantial reporting delays and large revisions indicate that the optimal Baseline specification may depend on revision patterns (Figure S1 and Figure S7). Although this may reflect features specific to the RSV time period examined, further evaluation using datasets with substantial retrospective revisions is needed to better understand Baseline behavior under such conditions. Finally, we did not evaluate more complex Baseline models, such as SARIMA or exponential smoothing approaches [32]. Our objective was to systematically examine how specification choices influence performance within simple, widely used Baseline models, and extending this analysis to more complex models that each had additional tuning parameters was beyond the scope of this study. Nevertheless, applying a similar sensitivity framework to commonly used complex Baseline models would be valuable for determining whether comparable generalizable performance patterns emerge.

In conclusion, our study demonstrates that specification choices within Baseline models can substantially influence forecast performance, model rankings, and the interpretation of comparative results. Across multiple respiratory disease targets, simple recent-data flatline Baseline models frequently achieved the strongest performance, even for diseases with pronounced seasonal dynamics. Nevertheless, our findings suggest that employing multiple Baseline models may be prudent for most forecasting hubs, providing complementary benchmarks for interpreting model performance. Most importantly, transparent documentation, consistent reporting, and clear archival records of Baseline configurations and updates over time are essential for ensuring reproducibility and interpretability of collaborative forecasting efforts.

## Data Availability

All data and code necessary to replicate the results presented within the study are available at
https://github.com/Ehsan-suez/baseline-work.

## Acknowledgements

We thank the attendees of the CSTE Forecasting All-Site Call, the EpiEngage All-Hands meetings, and the MIDAS Network Meeting for their helpful feedback and thoughtful discussions that informed this work. This work was supported in part by the Council of State and Territorial Epidemiologists (NU38OT000297); the Southwest Health Engagement and Research Collaborative (U54MD012388); the National Science Foundation (2230125); and the Centers for Disease Control and Prevention (75D30122C15411). This publication was additionally made possible by cooperative agreement CDC-RFA-FT-23-0069 from the CDC’s Center for Forecasting and Outbreak Analytics. The contents are solely the responsibility of the authors and do not necessarily represent the official views of the Centers for Disease Control and Prevention or other funders.

## Data and Code Availability

All data and code necessary to replicate the results presented within the study are available at https://github.com/Ehsan-suez/baseline-work.

