## Supplemental Information for "Basic Baseline model design choices can substantially influence performance in collaborative forecast hubs"

Ehsan Suez[1,2] and Spencer J. Fox[1,2,3]
[1] Institute of Bioinformatics, University of Georgia, Athens, GA, USA
[2] Center for the Ecology of Infectious Diseases, University of Georgia, Athens, GA, USA
[3] School of Informatics, Computing & Cyber Systems, Northern Arizona University, Flagstaff, AZ, USA

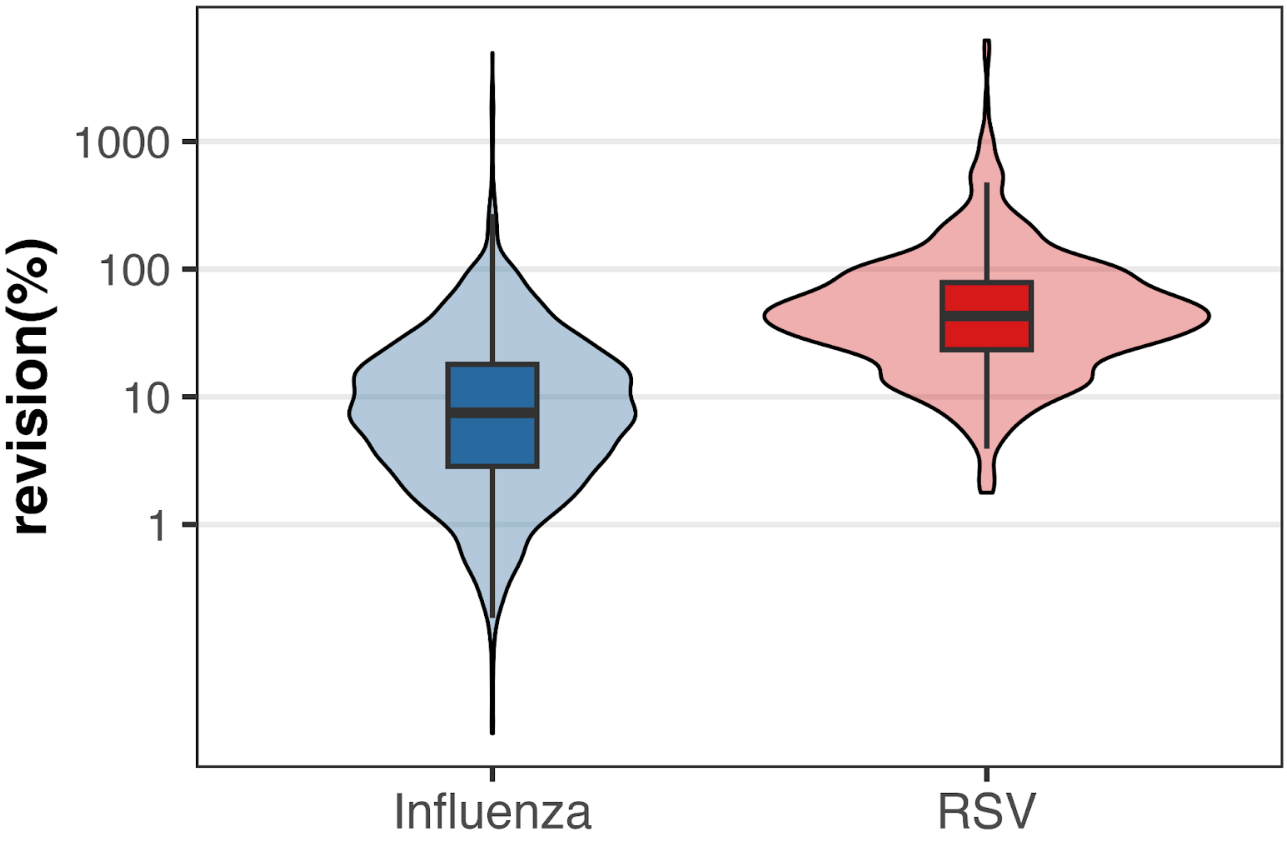


**Figure S1: Evaluation of data revisions for influenza and RSV hospital admissions.** Influenza hospital admissions are shown from October 2023 to May 2025, and RSV hospital admissions from January 2022 to April 2025, based on data available as of November 2025. Differences between initial and final reports reflect retrospective data revisions.

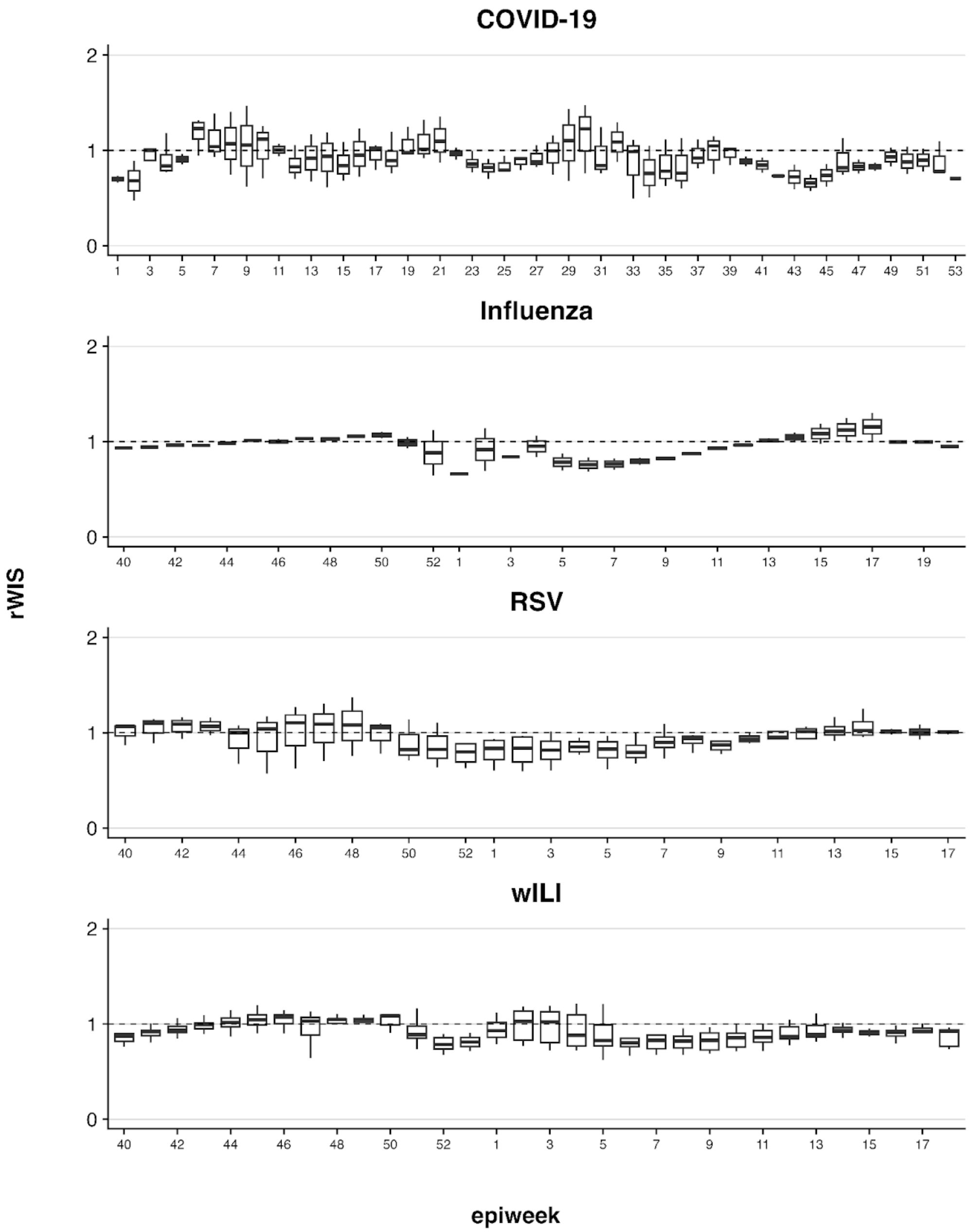


**Figure S2: Performance comparison between the drift and flatline Baseline models as a function of the epidemiological week across all regions, forecast horizons, and seasons under evaluation.** Weekly distributions of relative Weighted Interval Score (rWIS) are shown for COVID-19 (November 2020–October 2023), Influenza hospitalizations (October 2023-May 2025), RSV hospitalizations (January 2022-April 2025), and wILI (January 2014-April 2023) by epiweek. Boxplots summarize rWIS across all locations, forecast horizons, and forecast dates within each epidemiological week, with rWIS = WIS Flatline/ WIS Drift and values less than 1 indicating better performance of the flatline model. Horizontal dashed line indicates equal performance between the models.

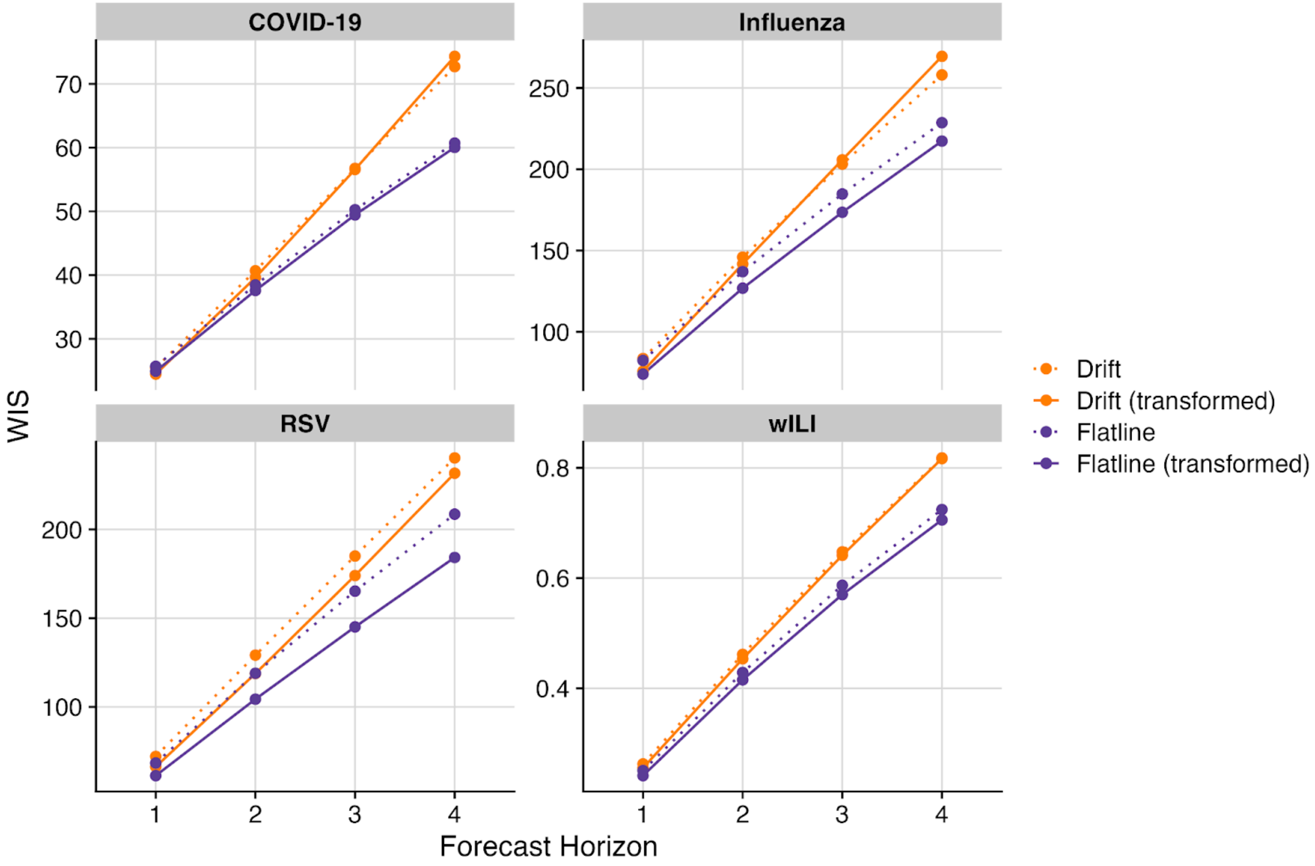


**Figure S3: Performance of the Baseline model variations as a function of the forecast horizon across all dates and regions.** WIS Mean Weighted Interval Score (WIS) is shown for Flatline and Drift baseline models with and without transformation for forecast horizons of 1-4 weeks ahead. Results are aggregated across all locations and forecast dates for COVID-19 (November 2020–October 2023), influenza hospitalizations (October 2023–May 2025), RSV hospitalizations (January 2022–April 2025), and wILI (January 2014–April 2023). Solid lines represent models fit on transformed data, while dotted lines represent models fit on the original scale. Lower WIS values indicate better probabilistic forecast performance. Lower WIS values indicate better probabilistic forecast performance.

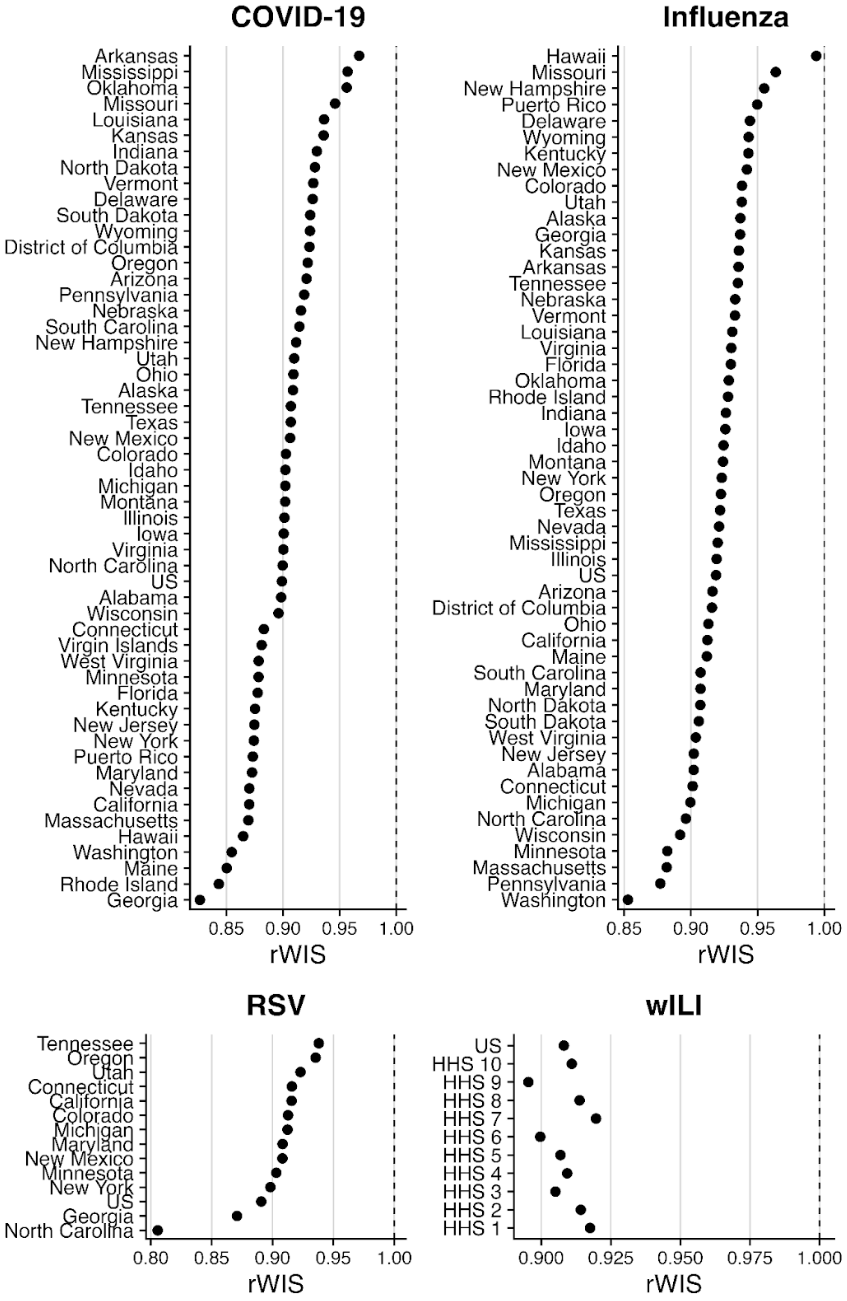


**Figure S4: Performance comparison between the flatline and drift model variants as a function of the forecast location.** Relative Weighted Interval Score (rWIS) is shown for each location for COVID-19, influenza hospitalizations, RSV hospitalizations, and wILI. Scores are aggregated across all forecast horizons and forecast dates within each evaluation period for COVID-19 (November 2020–October 2023), influenza hospitalizations (October 2023–May 2025), RSV hospitalizations (January 2022–April 2025), and wILI (January 2014–April 2023). The baseline models use the most recent 10 transformed observations for training. rWIS is defined as WIS Flatline/ WIS Drift, where values less than 1 indicate better performance of the flatline model. Each point corresponds to a forecast location (state, territory, or HHS region depending on the dataset), ordered by rWIS. The vertical dashed line at rWIS = 1 indicates equal performance between the two models.

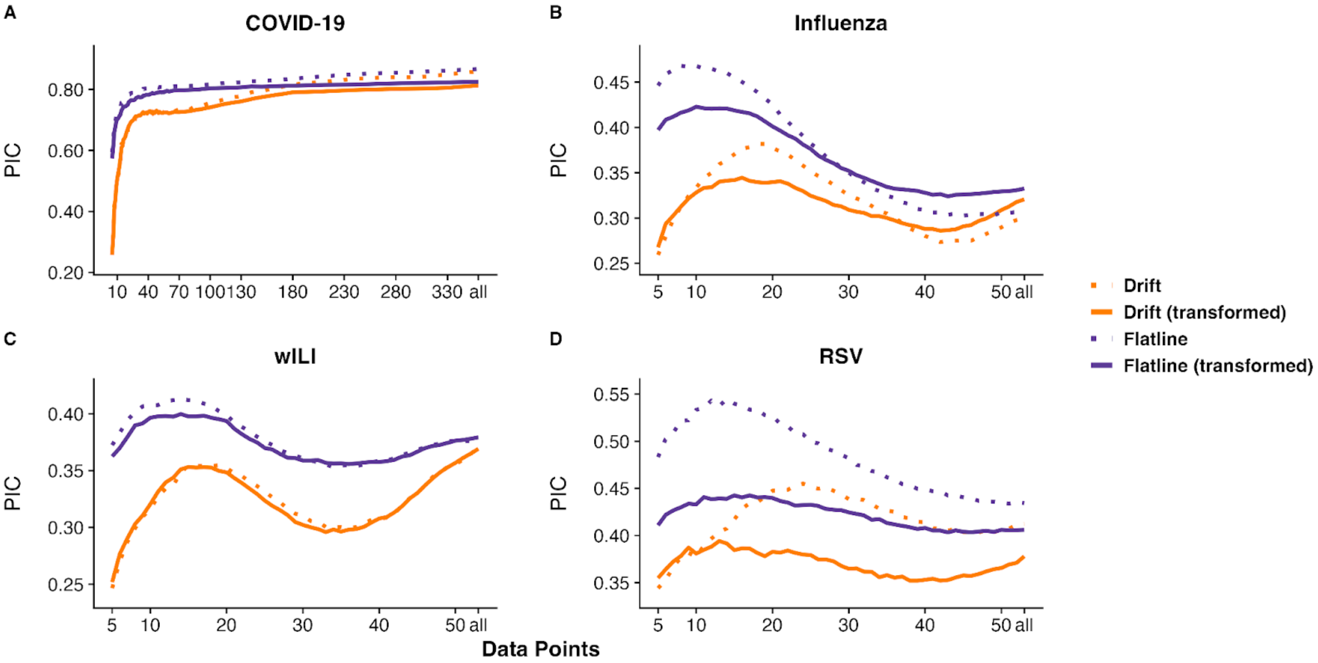


**Figure S5: Empirical 50% prediction interval coverage of baseline model variants as a function of the number of training observations.** Prediction interval coverage (PIC) for the empirical 50% prediction interval is shown as a function of the number of available training data points. Coverage is aggregated across all locations, forecast horizons, and forecast dates within each evaluation period for COVID-19 (November 2020–October 2023), influenza hospitalizations (October 2023–May 2025), RSV hospitalizations (January 2022–April 2025), and wILI (January 2014–April 2023). Panels A–D correspond to COVID-19, influenza hospitalizations, wILI, and RSV hospitalizations, respectively. Results are shown for Flatline and Drift baseline model variants with and without transformation. .

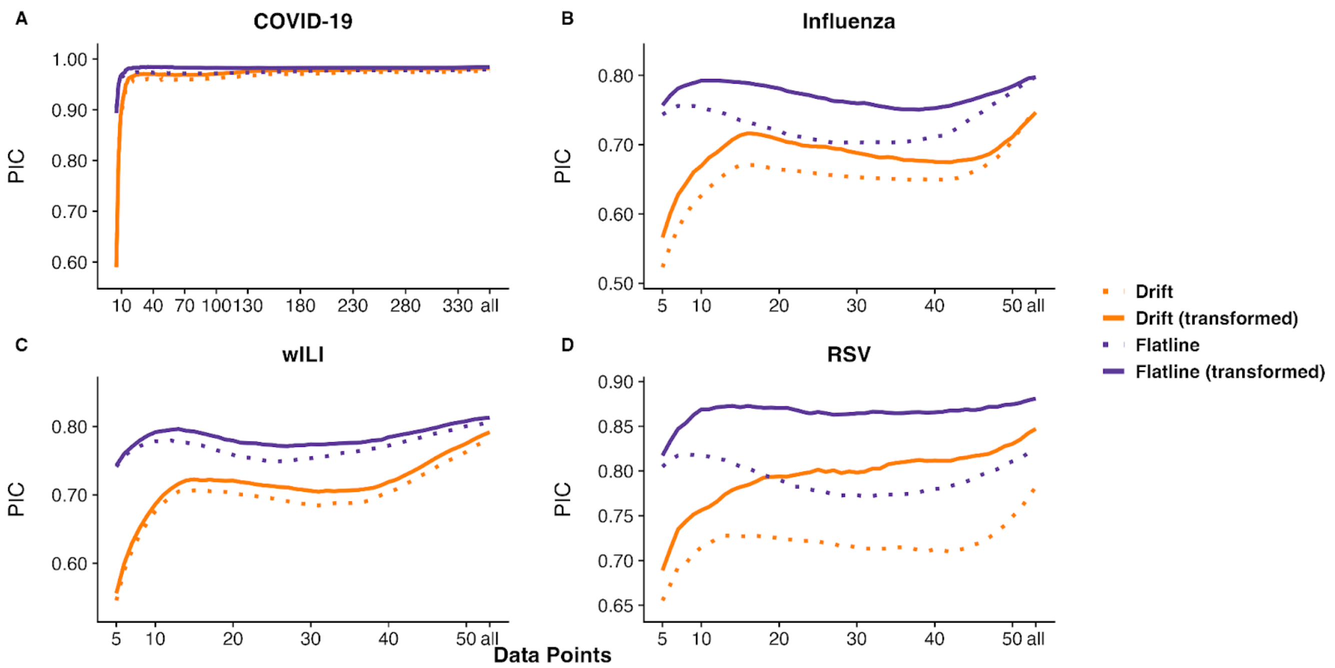


**Figure S6: Empirical 95% prediction interval coverage of baseline model variants as a function of the number of training observations.** Prediction interval coverage (PIC) for the empirical 95% prediction interval is shown as a function of the number of available training data points. Coverage is aggregated across all locations, forecast horizons, and forecast dates within each evaluation period for COVID-19 (November 2020–October 2023), influenza hospitalizations (October 2023–May 2025), RSV hospitalizations (January 2022–April 2025), and wILI (January 2014–April 2023). Panels A–D correspond to COVID-19, influenza hospitalizations, wILI, and RSV hospitalizations, respectively. Results are shown for Flatline and Drift baseline model variants with and without transformation.  .

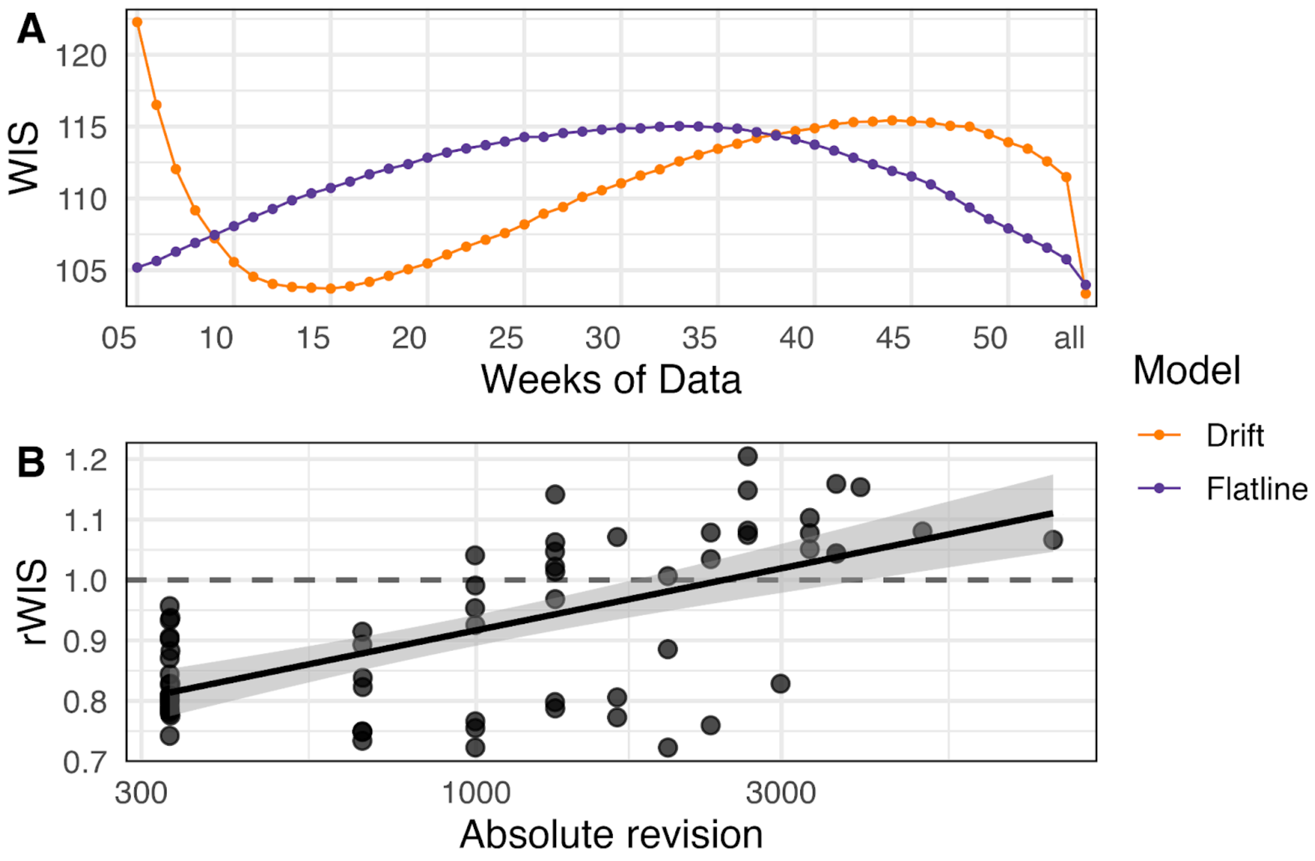


**Figure S7: Sensitivity of RSV baseline performance to reporting delays.**
(**A**) Mean weighted interval score (WIS) for Drift and Flatline baseline models as a function of the number of weeks of training data when used as-of data. (**B**) Relationship between reporting revision magnitude and relative forecast performance (rWIS = WIS Flatline / WIS Drift). Each point represents a forecast date. The dashed line indicates equal performance (rWIS = 1); values below this line indicate better performance of the Flatline model, while values above indicate better performance of the Drift model.

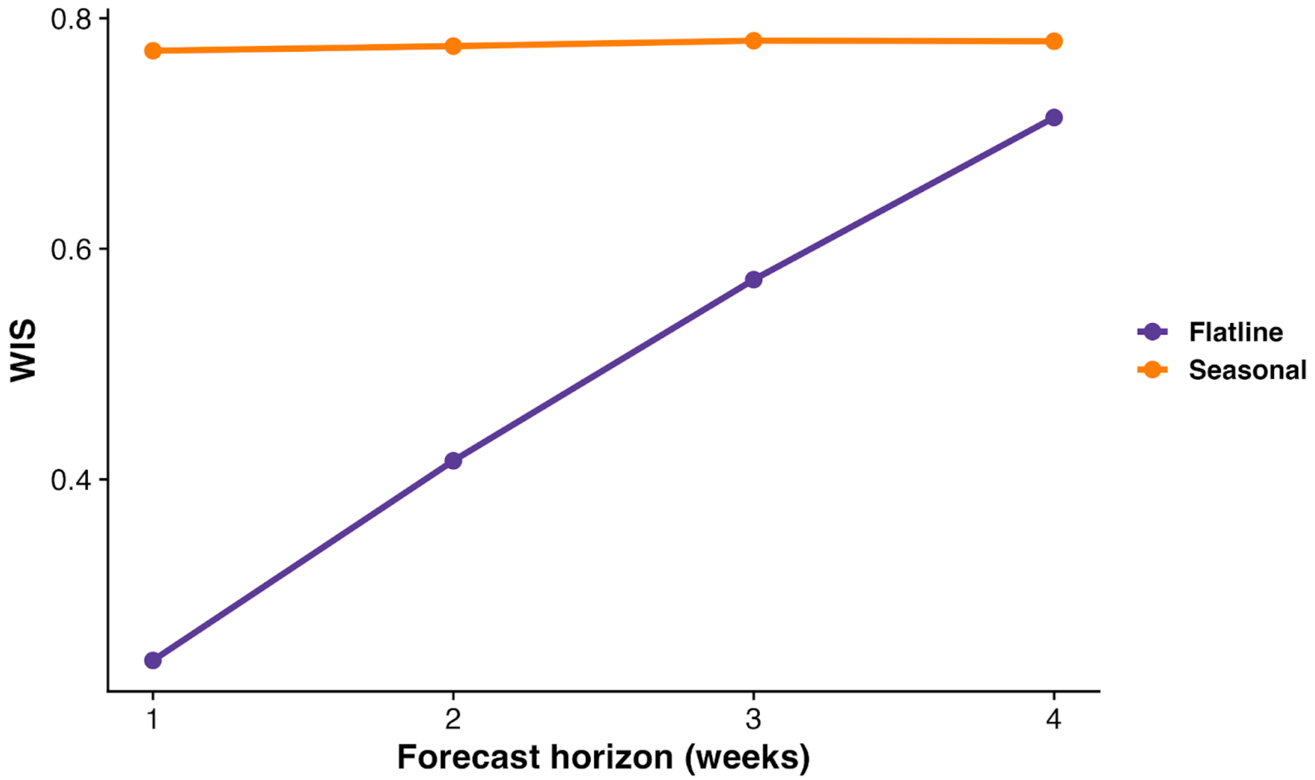


**Figure S8: Performance comparison between the Flatline and Seasonal baseline models as a function of forecast horizon.** Mean Weighted Interval Score (WIS) is shown for the Flatline and Seasonal baseline models for forecast horizons of 1–4 weeks ahead. Scores are aggregated across all locations and forecast dates within the evaluation period for wILI (January 2014–April 2023). Lines represent the average WIS for each model at each forecast horizon. Lower WIS values indicate better probabilistic forecast performance. .

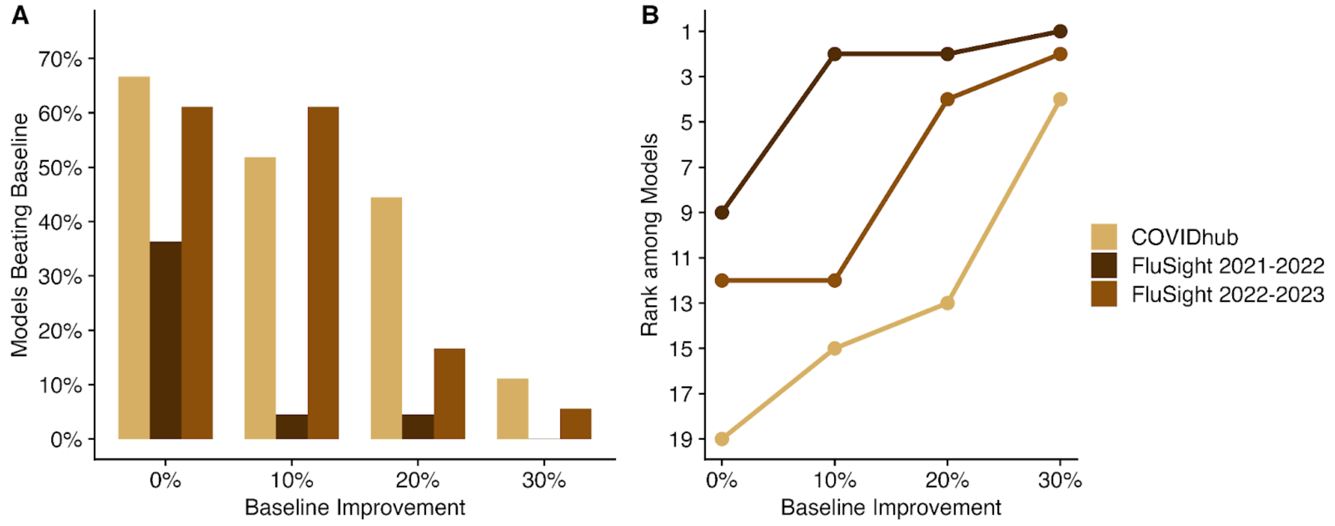


**Figure S9:Potential impact of improving the Baseline model performance on relative model performance from recent real-time forecast efforts. (A)** Percentage of models that would outperform the Baseline model for the COVID-19 forecast hub and the 2021-2022 and 2022-2023 FluSight challenges (colored bars) if the Baseline model performance improved by a specified percentage. **(B)** Baseline model rank across all submitted forecast models if its performance improved by a specified percentage for the COVID-19 forecast hub and the 2021-2022 and 2022-2023 FluSight challenges (colored points and lines). Forecasts were evaluated from April 2020 through October 2021 for COVID-19, and respective influenza seasons for 2021-2022 FluSight and 2022-2023 FluSight.
